# The individual and combined associations of depression and socioeconomic status with risk of major cardiovascular events: a prospective cohort study

**DOI:** 10.1101/2021.12.31.21268566

**Authors:** Regina Prigge, Sarah H. Wild, Caroline A. Jackson

## Abstract

**Objective:** We aimed to investigate the individual and combined associations of depression and low socioeconomic status (SES) with risk of major cardiovascular events (MCVE), defined as first-ever fatal or non-fatal stroke or myocardial infarction, in a large prospective cohort study.

**Methods:** We used data from 466,238 UK Biobank participants, aged 40 – 69 years without cardiovascular disease, bipolar disorder or schizophrenia at baseline. We performed Cox proportional hazard models to estimate adjusted hazard ratios (HR) and 95% confidence intervals (CI) of the individual and combined associations of depression and each of educational attainment, area-based deprivation and income with risk of MCVE. We assessed effect modification and explored interaction on the additive and multiplicative scale.

**Results:** Depression, low education, high area-based deprivation and low income were individually associated with increased risks of MCVE (adjusted HR, 95% CI: 1.28, 1.19 – 1.38; 1.20, 1.14 – 1.27; 1.17, 1.11 – 1.23; and 1.22, 1.16 – 1.29, respectively). Depression was associated with increased risks of MCVE among individuals with high and low SES. Individuals with depression and each of low education, high area-based deprivation and low income were at particularly high risk of MCVE (HR, 95% CI: 1.50, 1.38 – 1.63; 1.63, 1.46 – 1.82; 1.31, 1.23 – 1.40, respectively). There was interaction between depression and area-based deprivation on multiplicative and additive scales but no interaction with education or income.

**Conclusion:** Depression was associated with increased risks of MCVE among individuals with high and low SES, with particularly high risks among those living in areas of high deprivation.

## BACKGROUND

Depression and low socioeconomic status (SES) are both associated with increased risks of physical diseases, such as myocardial infarction (MI) and stroke [1-6]. Furthermore, individuals with a low SES are disproportionally affected by mental-physical comorbidity from early adulthood onwards [7, 8] with depression being the most common mental health condition [9]. It has been hypothesized that individuals with a low SES are exposed to more adversity whilst having fewer resources to cope with these stressors [10, 11]. Thus, individuals with a low SES may be particularly vulnerable to the adverse effects of depression on risk of major cardiovascular events (MCVE).

There is growing evidence that the association between depression and all-cause and cardiovascular mortality is more pronounced among individuals with a low SES [11, 12]. Less information is available on the association between depression and incident cardiovascular disease (CVD), with existing studies providing inconsistent results. Whilst some studies have shown that individuals with a low SES might be more vulnerable to the adverse effects of depression than those with a high SES for one or both of MI and stroke [13-16], other studies have reported that individuals with a high SES may be more vulnerable [17], or have not observed any differences between groups [18-21]. Potential reasons for the conflicting findings are the use of different indicators of SES, the use of different methodological approaches to investigate effect modification and/or interaction, and differences in the study populations. The inconsistent effects of different SES measure is highlighted by a US-based study on the association between depression and incident coronary heart disease or revascularization that reported effect modification by income but not education [14].

Due to the inconsistent findings and methodological shortcomings of existing studies, a recent paper highlighted the need for further studies investigating the individual and combined effect of depression and SES on incidence of cardiovascular events [22]. Advancing in our understanding of the relationship between depression, SES and risk of MCVE might allow the identification of individuals that are particularly vulnerable to the adverse effects of depression which in turn might aid tailoring of resources and preventive measures. Furthermore, it might help inform our understanding of the underlying mechanisms of the association between depression and MCVE. Thus, we aimed to investigate the individual and combined associations of depression and different measures of SES with risk of MCVE in a large prospective cohort study in the United Kingdom.

## METHODS

### Study population

The UK Biobank is a prospective cohort study of ∼500,000 participants who were recruited in England, Scotland, and Wales from 2006 to 2010. Participants were aged 40 – 69 years at baseline. We excluded participants who withdrew from the study, whose information could not be linked to hospital or death records, and who had a history of CVD (defined as stroke, MI, angina or transient ischaemic attack), bipolar disorder or schizophrenia at baseline. Participants were followed up through linkage to routinely collected health and death records [23]. Analyses of UK Biobank data are conducted under generic approval from the NHS National Research Ethics Service (Ref 11/NW/0382, approval letter dated 17 June 2011). Full written informed consent was obtained from participants at the point of data collection.

### Exposure

We defined depression as at least one of: self-report of depression or antidepressant use; or hospital record of depression at baseline. We identified self-reported antidepressant use and self-reported doctor-diagnosis of depression through information obtained in the nurse interview at baseline. Antidepressant use was defined as self-reported use of at least one selective serotonin reuptake inhibitor or other antidepressant medication [24]. We identified a hospital record of depression using ICD 10 codes F32 – F33. We used educational attainment, area-based deprivation and income as measures of SES, all of which were treated as binary variables based on information collected in a touchscreen questionnaire at baseline. We defined educational attainment as high and low based on the presence or absence of a university or college degree, respectively. Income was treated as low if the annual household income was less than £31,000 or high if the annual household income was greater or equal to £31,000, which was close to the median disposable household income in the UK in 2020 [25]. Area-based deprivation was defined using the Townsend index [26]. A score of 0 represented an area in the United Kingdom with average area-based deprivation. We defined areas with high deprivation as those with positive scores and areas with low deprivation as those with negative scores.

### Outcome

MCVE were defined as first-ever fatal or non-fatal stroke or MI, whichever occurred first. We used linkage to hospital inpatient and death records to identify fatal and non-fatal stroke and MI during follow-up [27]. We defined stroke using ICD 10 codes I60, I61, I63 and I64 and MI using I21, I22, I24.1 and I25.2. Survival times were calculated from the date of attending the baseline assessment centre to the date of first-ever MCVE, date of death, or end of follow-up (31 March 2015).

### Covariates

Covariates included age, sex, ethnicity, body mass index, smoking, alcohol intake, physical activity, fruit and vegetable intake, hypertension, diabetes, high cholesterol, family history of CVD, and family history of severe depression. Information on these variables were ascertained through self-report in the touchscreen questionnaire and nurse interview and, where available, through measured values at baseline or hospital records prior to baseline of the study (Supplementary material S1).

### Statistical analysis

We performed analyses using R version 4.0.0. We compared baseline characteristics of participants with and without MCVE during follow-up. We used Cox proportional hazards models to estimate hazard ratios (HR) and 95% confidence intervals (CI) of the individual and combined associations of depression and each SES measure on the risk of MCVE. Individual associations were assessed in an unadjusted model, a model that was adjusted for age, sex and ethnicity, and a third model that additionally controlled for body mass index, alcohol intake, physical activity, smoking, fruit and vegetable consumption, oily fish intake, history of hypertension, diabetes, and high cholesterol levels, and family history of CVD or depression. We then assessed the combined association of depression and each SES measure on risk of MCVE in the fully adjusted model. We performed pre-specified sex-stratified analyses to evaluate differences between men and women. We tested the proportional hazards assumption for all variables using the cox.zph function, and investigated potential violations using log-minus-log survival plots and plots of scaled Schoenfeld residuals against time. The proportional hazards assumption was met for all variables.

As recommended by Sullivan and Vaccarino [22], we tested for effect modification, additive and multiplicative interaction, following established reporting guidelines for these measures [28]. The effect modification and interaction analyses were performed in keeping with the code of the interactionR package [29]. Accompanying 95% CIs were calculated using the delta method [30]. We investigated for additive (i.e. biological) interaction by calculating the relative excess risk due to interaction (RERI). Multiplicative interaction was investgated by adding a product term of depression and each SES measure to the fully adjusted Cox proportional hazards model. We considered a two-sided p<0.05 statistically significant.

We determined a a violation of the missing completely at random (MCAR) assumption since there were differences between participants with and without complete data (Supplementary material S2). Due to the large number of included variables, 143,368 (30.7%) participants had at least one missing value in any variable. Since a missing at random mechanism was deemed likely, multiple imputation of missing data was performed using the MICE package in R [31]. In keeping with the recommendation that the number of imputations should equate to the percent of incomplete cases [32], we performed 32 imputations with 10 iterations. Since we were interested in interaction effects, we computed imputations separately for participants with and without depression. Since results of complete cases analyses are potentially biased when data are not MCAR, we present results based on imputed data, with results of the complete case analysis provided in Supplementary material S3.

### Sensitivity analysis

First, since existing studies reported varying results for different components of our primary outcome [15, 16], we repeated the analysis for stroke and MI separately. Second, since competing risks during follow-up may have introduced bias, we investigated to what extent deaths from causes other than stroke and MI might affect associations between depression and MCVE by running a competing risk analysis.

## RESULTS

### Descriptive statistics

After excluding people with CVD, schizophrenia or bipolar disorder at baseline, we included 466,238 people with a median age of 57 years (IQR: 50 – 63 years) in our analysis (Figure 1). Of these, 205,902 (44.2%) were male, 40,649 (8.7%) were categorised as having depression, 303,605 (65.1%) had low educational attainment, 131,037 (28.1%) lived in areas with high deprivation, and 184,786 (39.6%) had low income at baseline.

**Figure 1.**
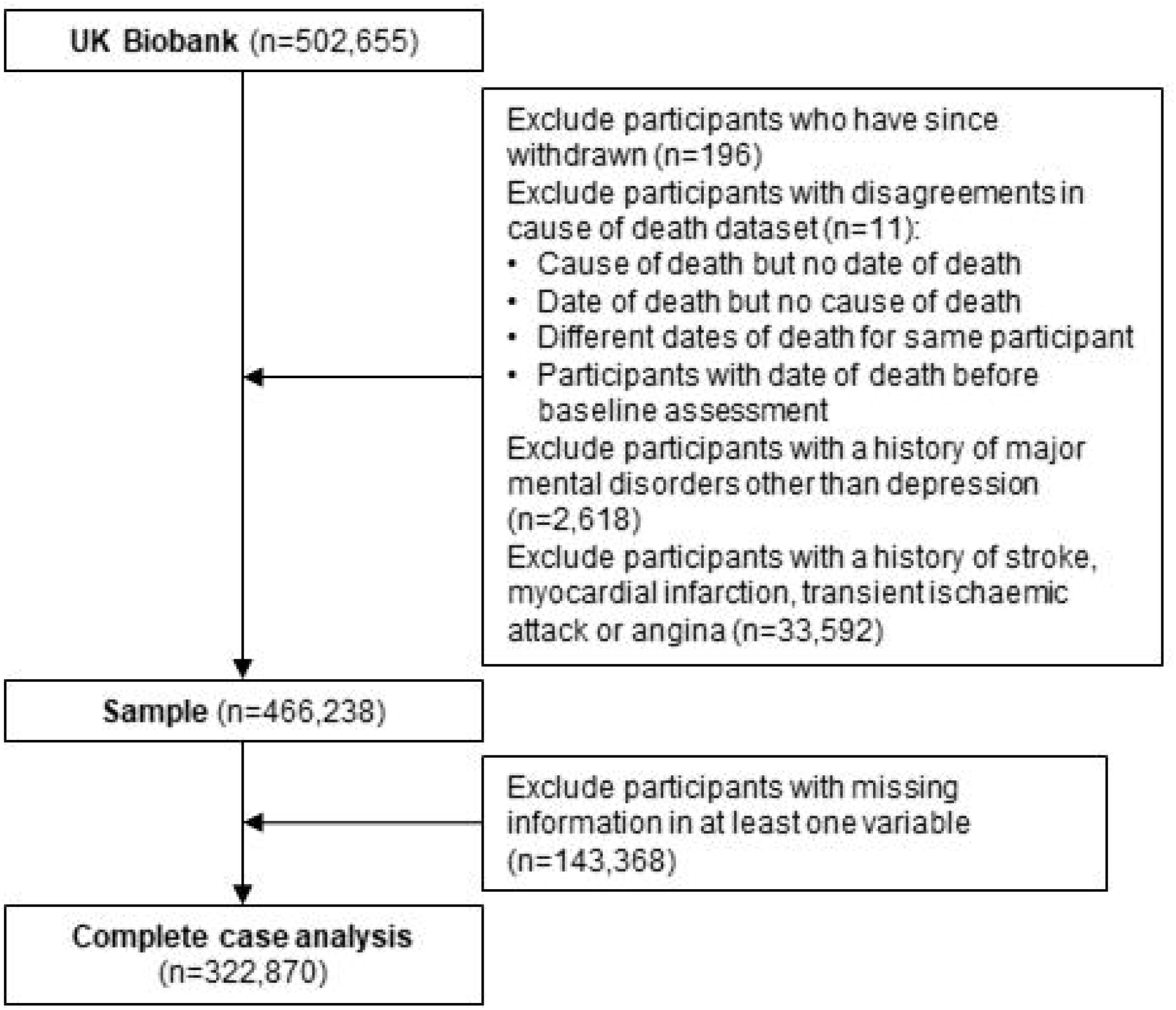
Flow diagram of sample selection

There were 7,675 fatal or non-fatal MCVE during a median of 6.1 (IQR: 5.4 – 6.8) years of follow-up (Table 1). At baseline, the prevalence of depression, low educational attainment, high area-based deprivation and low income was higher among those with MCVE than among those with no events during follow-up. Furthermore, those who developed MCVE during follow-up reported higher proportions of hypertension, diabetes, high cholesterol, overweight or obesity, and smoking at baseline. The proportion with MCVE was lowest among individuals without depression and each of high education, low area-based deprivation and high income, and highest among those with depression and low education, depression and high area-based deprivation, and those with low income in the absence of depression.

**Table 1:**
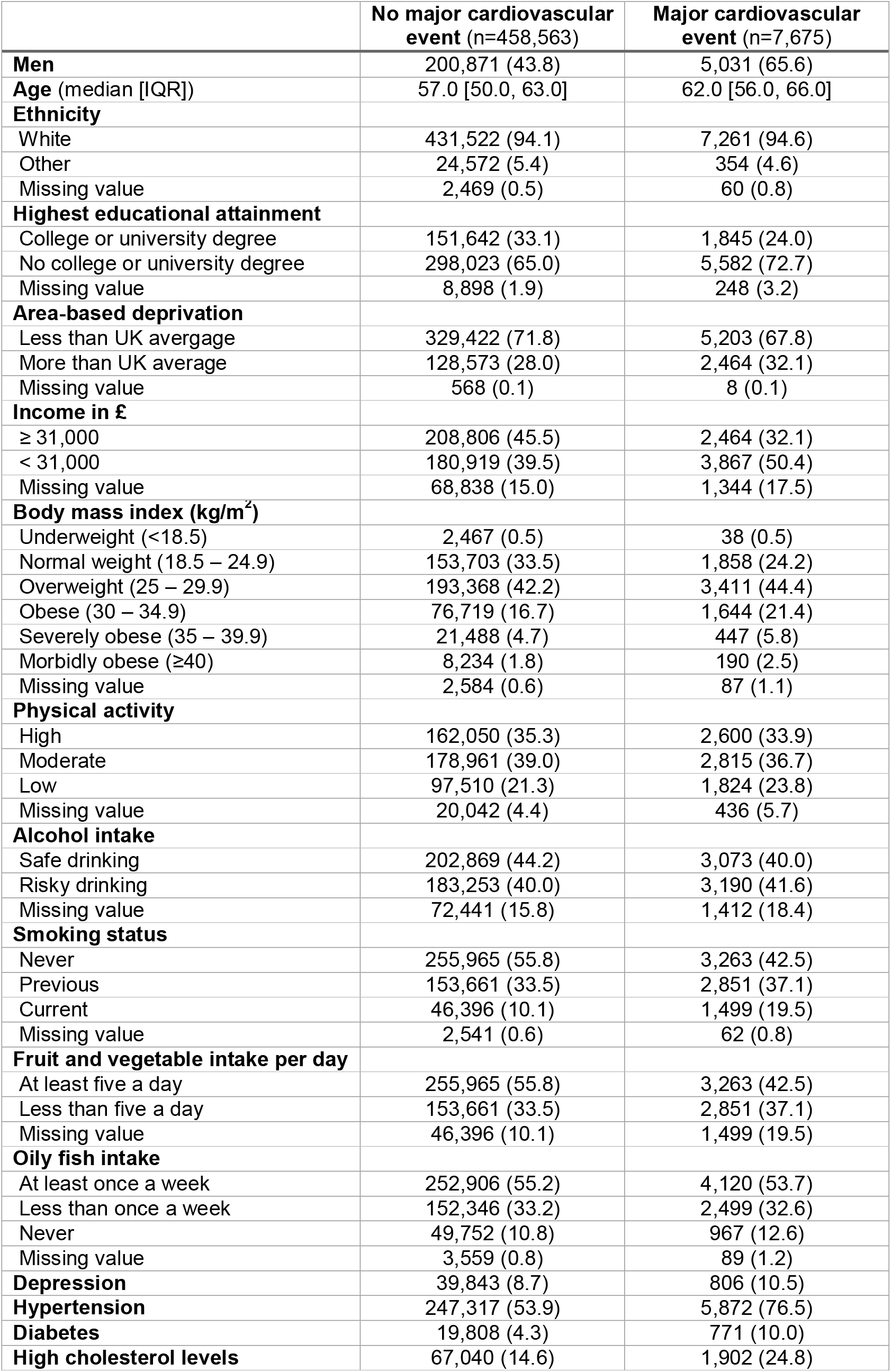
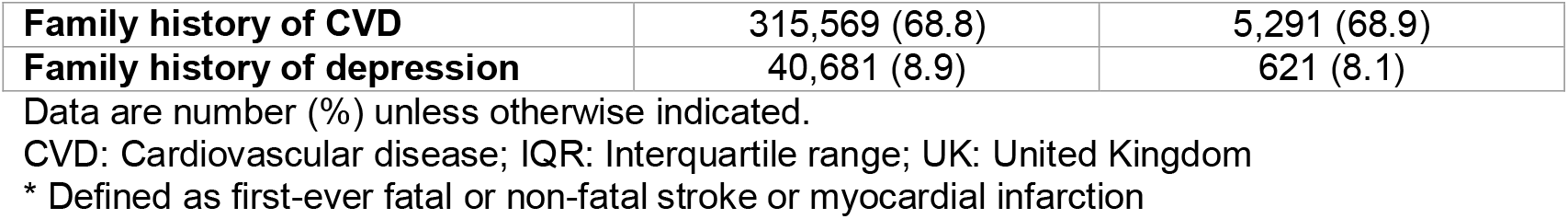
Baseline characteristics and for UK Biobank participants with and without incident major cardiovascular events* during follow-up

### Individual associations of depression and SES measures with risk of MCVEs

Each of depression, low education, high area-based deprivation and low income were individually associated with increased risks of MCVE in unadjusted, partially adjusted and fully adjusted models (Table 2). The associations attenuated but remained statistically significant after adjusting for various factors. After adjustment for all covariates (Model 3), depression was associated with a 28% increased risk of MCVE (HR, 95% CI: 1.28, 1.19 – 1.38), and education, area-based deprivation and income were associated with 20%, 17% and 23% increased risks of MCVE, respectively (HR, 95% CI: 1.20, 1.14 – 1.27; 1.17, 1.11 – 1.23; and 1.22, 1.16 – 1.29, respectively). Mutual adjustment for depression and each of education, deprivation and income changed the effect estimates marginally.

**Table 2:** Hazard ratios (95% CI) of the individuals effects of depression and socioeconomic status on risk of major cardiovascular events*

|  | **Depression**<br>(yes vs no) | **Educational attainment**<br>(low vs high) | **Area-based deprivation**<br>(high vs low) | **Income**<br>(low vs high) |
| --- | --- | --- | --- | --- |
| **Model 1** | 1.23 (1.14 – 1.32) | 1.53 (1.45 – 1.61) | 1.24 (1.18 – 1.30) | 1.79 (1.70 – 1.88) |
| **Model 2** | 1.48 (1.37 – 1.59) | 1.40 (1.33 – 1.47) | 1.35 (1.29 – 1.42) | 1.40 (1.33 – 1.48) |
| **Model 3** | 1.28 (1.19 – 1.38) | 1.20 (1.14 – 1.27) | 1.17 (1.11 – 1.23) | 1.22 (1.16 – 1.29) |
| **+ depression** | — | 1.20 (1.14 – 1.27) | 1.16 (1.10 – 1.22) | 1.21 (1.15 – 1.28) |
| **+ education** | 1.27 (1.18 – 1.37) | — | — | — |
| **+ deprivation** | 1.27 (1.18 – 1.37) | — | — | — |
| **+ income** | 1.25 (1.16 – 1.35) | — | — | — |

Data are hazard ratios (95% confidence intervals)
\*Model 1: Depression, socioeconomic factor, age, sex, ethnicity
†Model 2: Model 1 + body mass index, physical activity, alcohol intake, smoking, fruit and vegetable intake, oily fish intake, high cholesterol levels, hypertension, diabetes, family history of cardiovascular disease, and family history of depression
\* Defined as first-ever fatal or non-fatal stroke or myocardial infarction

### Combined associations of depression and SES measures with risk of MCVEs

Irrespective of the SES measure used, depression was associated with increased risk of MCVE in low and high SES categories, and low SES was associated with increased risks of MCVE among people with and without depression (Table 3). Whilst the risk of MCVE associated with depression was similar among people with low and high education (HR, 95% CI: 1.28, 1.17 – 1.39 and 1.23, 1.05 – 1.45, respectively) and people with low and high income (HR, 95% CI: 1.26, 1.15 – 1.37 and 1.18, 1.01 – 1.39, respectively), the risk of MCVE among people with depression was higher in individuals living in areas of high deprivation than in people living in areas with low deprivation (HR, 95% CI: 1.42, 1.27 – 1.60 and 1.14, 1.04 – 1.26, respectively).

**Table 3:**
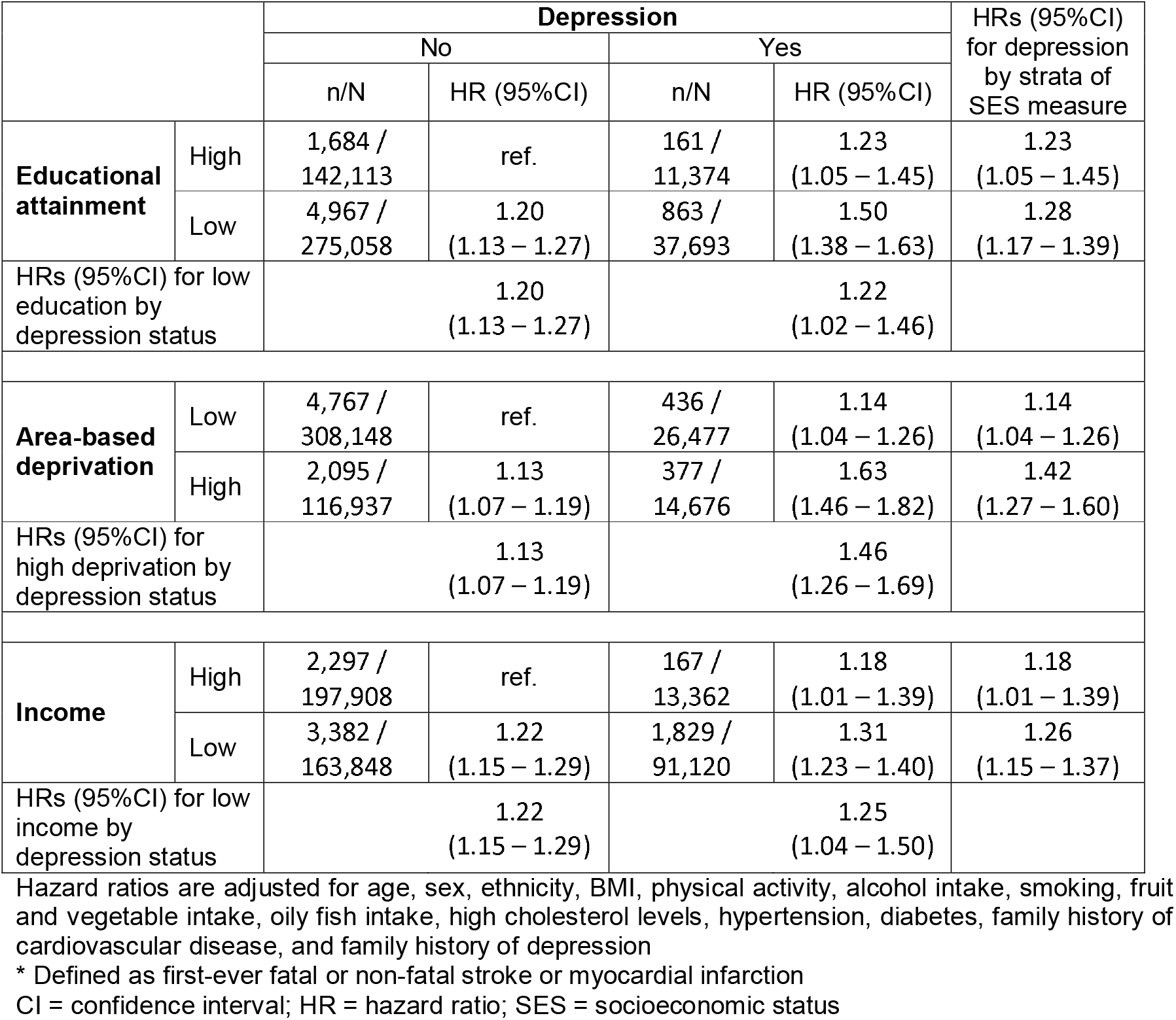
Numbers of events and hazard ratios (95% CI) for the combined association of depression and different measures of socioeconomic status on risk of major cardiovascular events*

|  |  | Depression |  |  |  | HRs (95%CI)<br>for depression<br>by strata of<br>SES measure |
| --- | --- | --- | --- | --- | --- | --- |
|  |  | No |  | Yes |  |  |
|  |  | n/N | HR (95%CI) | n/N | HR (95%CI) |  |
| Educational attainment | High | 1,684 /<br>142,113 | ref. | 161 /<br>11,374 | 1.23<br>(1.05 – 1.45) | 1.23<br>(1.05 – 1.45) |
|  | Low | 4,967 /<br>275,058 | 1.20<br>(1.13 – 1.27) | 863 /<br>37,693 | 1.50<br>(1.38 – 1.63) | 1.28<br>(1.17 – 1.39) |
| HRs (95%CI) for low education by depression status |  | 1.20<br>(1.13 – 1.27) |  | 1.22<br>(1.02 – 1.46) |  |  |
| Area-based deprivation | Low | 4,767 /<br>308,148 | ref. | 436 /<br>26,477 | 1.14<br>(1.04 – 1.26) | 1.14<br>(1.04 – 1.26) |
|  | High | 2,095 /<br>116,937 | 1.13<br>(1.07 – 1.19) | 377 /<br>14,676 | 1.63<br>(1.46 – 1.82) | 1.42<br>(1.27 – 1.60) |
| HRs (95%CI) for high deprivation by depression status |  | 1.13<br>(1.07 – 1.19) |  | 1.46<br>(1.26 – 1.69) |  |  |
| Income | High | 2,297 /<br>197,908 | ref. | 167 /<br>13,362 | 1.18<br>(1.01 – 1.39) | 1.18<br>(1.01 – 1.39) |
|  | Low | 3,382 /<br>163,848 | 1.22<br>(1.15 – 1.29) | 1,829 /<br>91,120 | 1.31<br>(1.23 – 1.40) | 1.26<br>(1.15 – 1.37) |
| HRs (95%CI) for low income by depression status |  | 1.22<br>(1.15 – 1.29) |  | 1.25<br>(1.04 – 1.50) |  |  |

Hazard ratios are adjusted for age, sex, ethnicity, BMI, physical activity, alcohol intake, smoking, fruit and vegetable intake, oily fish intake, high cholesterol levels, hypertension, diabetes, family history of cardiovascular disease, and family history of depression
\* Defined as first-ever fatal or non-fatal stroke or myocardial infarction
CI = confidence interval; HR = hazard ratio; SES = socioeconomic status

Depression alone, low socioeconomic status alone and combined depression and low socioeconomic status were associated with greater risks of MCVE compared to those with no depression and high SES, irrespective of the SES measure used (Table 3). For each socioeconomic measure the associations followed similar patterns, with similar risks of MCVE among those with depression alone and low SES alone and highest effect estimates for those with both depression and low SES, compared to those with no depression and high SES. The combined associations of depression and each of low education, high area-based deprivation, and low income on risk of MCVE were 1.50 (1.38 – 1.63), 1.63 (1.46 – 1.82), and 1.31 (1.23 – 1.40), respectively, compared to those with no depression and high SES.

### Additive and multiplicative interaction

There was multiplicative interaction between depression and area-based deprivation (p<0.001), with the effect of depression significantly stronger among those living in areas with high deprivation, but no multiplicative interaction for depression and either education or income (p = 0.667 and p = 0.550, respectively) (Table 4). The estimated combined effects of depression and area-based deprivation exceeded the sum of the risk of depression alone and area-based deprivation alone, indicating evidence of interaction on the additive scale (RERI: 0.36, 95% CI: 0.15 – 0.56, p<0.001). However, there was no evidence of additive interaction between depression and education (RERI: 0.07, 95% CI: -0.15 – 0.29, p=0.267) or depression and income (RERI: -0.09, 95% CI: -0.29 – 0.11, p=0.799).

**Table 4:** Measures of additive and multiplicative interaction between depression and different measures of socioeconomic status on risk of major cardiovascular events*

|  | **Additive interaction**<br>(RERI, 95% CI, p-value) | **Multiplicative interaction**<br>(Ratio of HRs for SES measure<br>within strata of depression,<br>95% C, p-value) |
| --- | --- | --- |
| **Depression and education** | 0.07 (-0.15 – 0.29), p=0.267 | 1.04 (0.87 – 1.25), p=0.667 |
| **Depression and area-based deprivation** | 0.36 (0.15 – 0.56), p<0.001 | 1.28 (1.10 – 1.48), p=0.001 |
| **Depression and income** | -0.09 (-0.29 – 0.11), p=0.799 | 1.06 (0.88 – 1.27), p=0.559 |

Estimates (95% CI) are based on Cox proportional hazards models adjusted for age, sex, ethnicity, BMI, physical activity, alcohol intake, smoking, fruit and vegetable intake, oily fish intake, high cholesterol levels, hypertension, diabetes, family history of cardiovascular disease, and family history of depression
\* First-ever fatal or non-fatal stroke or myocardial infarction
CI = confidence interval; HR = hazard ratio; RERI = Relative Excess Risk due to Interaction; SES = socioeconomic status

### Sex-stratified analysis

Each of depression, education, deprivation, and income were individually associated with risk of MCVE in men and women (Supplementary material S4). The strength of the individual association between each of educational attainment and income and risk of MCVE was similar for men and women, whereas the associations of depression and area-based deprivation on risk of MCVE was stronger among women than men. In each stratum of high and low SES, the relative risks of MCVE associated with depression were higher among women than men. Irrespective of the SES measure used, the risk of MCVE was highest among men and women with depression and a low SES, compared to those with no depression and a high SES, with the exception that the risk of women with depression alone exceeded the risk of women with depression and low income. However, confidence intervals were wide and overlapped. There was no evidence of interaction between depression and each of education and income for men and women, whereas there was evidence of interaction between depression and area-based deprivation on the additive and multiplicative scale for women but not for men.

### Sensitivity analyses

Depression, education, area-based deprivation and income were individually associated with increased risks of each of stroke and MI (Supplementary material S5). Depression was generally associated with increased risks of stroke and MI among people with low and high SES, irrespective of the measure used. However, some estimates were imprecise. Furthermore, those with depression and low SES were at highest risk of both stroke and MI, compared to people with no depression and high SES. In line with the findings of our primary analysis, we found additive interaction between depression and area-based deprivation, but not education and income, when we analysed MI and stroke separately.

Each of depression, education, area-based deprivation and income were individually associated with risk of death due to causes other than stroke or MI (Supplementary material S6). Depression was associated with increased risk of competing events among those with low and high SES, irrespective of the SES measures used. Furthermore, those with combined depression and one of low education, high area-based deprivation and low income were at highest risk of the competing event, relative to those with no depression and high SES. There was no evidence for interaction between depression and any SES measure on the additive or multiplicative scale for competing risks.

## DISCUSSION

In a large prospective cohort study in the United Kingdom, each of depression, low education, high area-based deprivation and low income were individually associated with risk of MCVE. Furthermore, depression was associated with increased risk of MCVE among people with low and high SES, irrespective of which measure of SES was used. Whilst we did not find any interaction between depression and either education or income, we did find interaction between depression and area-based deprivation on the additive and multiplicative scales that was more marked among women. Competing events did not explain the observed associations.

### Strength and limitations

Our study has a number of strengths. To our knowledge, this is the most comprehensive assessment of the individual and combined associations of depression and SES on risk of MCVE using different measures of SES and following recommended reporting guidelines [28]. Our analysis was sufficiently powered to precisely estimate the individual and combined associations of depression and different measures of SES on risk of MCVE due to the large sample size and number of MCVE in UK Biobank. Furthermore, we were able to stratify our analysis by sex and adjust our effect estimates for various demographic and lifestyle factors due to the large amount of information collected at the UK Biobank baseline assessment. Since we used administrative health records to ascertain outcomes, we had limited loss to follow-up.

Our study has some limitations. First, the low response rate of UK Biobank (5.5%) may have introduced bias. Whilst a study population that is composed of relatively healthy individuals from higher than average or median socioeconomic backgrounds has obvious implications on estimating prevalence and incidence, it has been argued that selection does not influence associations between baseline characteristics and health outcomes due to the large number of individuals with different levels of socioeconomic factors [33]. Second, whilst record linkage to hospital and death records offered many benefits, we will have missed people with non-fatal events who were not admitted to hospital. However, systematic reviews on the accuracy of hospital and death records for identifying events of stroke and MI concluded that positive predictive values of more than 90% can be achieved when stroke-specific codes were chosen and hospital records were used to detect MI [34, 35]. Third, there was potential for misclassification of the exposure status of individuals at baseline. Despite using self-report and hospital records to identify individuals with depression at baseline, depression may have been underreported at baseline which may have diluted our effect estimates. Furthermore, there is potential for misclassification of levels of SES. Common limitations in the measurement of SES have been widely described [36], such as high rates of non-response or misclassification of individuals to SES levels based on area-based SES measures. We have also only used binary categories of SES. Fourth, despite controlling for various potential confounding factors in our analysis, our results may be affected by residual confounding due to measurement error of included covariates or unmeasured covariates.

### Comparison with previous studies

Our results are in keeping with previous studies highlighting a high risk of CVD associated with each of depression and low SES individually [1-6, 13, 15]. As highlighted by Sullivan and Vaccarino [22], a comparison of findings of existing studies on the combined association of depression and SES on risk of CVD is complicated since these studies did not present all information required to assess effect modification and interaction [28]. Nonetheless, our results are largely consistent with existing studies. The most comprehensive existing study of individuals from South Korea found that depression was associated with increased risks of stroke and MI among those with high and low insurance premiums (used as a proxy measure of income) [15]. Furthermore, those with low income and depression had the highest risk of stroke and MI compared to those with no depression and high income. There was no evidence of multiplicative interaction and authors concluded that depression and income had additive effects on risk of stroke and MI, which is in keeping with our findings on income. In keeping with our study, existing studies generally did not find interaction between depression and each of education, income and receipt of social security assistance on risk of stroke or IHD on the multiplicative scale [18, 19, 21]. In contrast to our findings, depressive symptoms were associated with incident CHD or revascularization among individuals with low income but not among those with high income in a US-based study [14]. However, differences are perhaps expected given the universal access to health care is available in the UK and not in the US. To our knowledge, there is only one existing study assessing the combined effect of depression or bipolar disorder and area-based deprivation on the incidence of IHD and stroke [16]. Similar to our study, there was a suggestion of interaction on the additive scale on risk of IHD, with the risk most pronounced among the most deprived groups. However, in contrast to our findings, the authors found no interaction between depression and area-based deprivation on risk of stroke.

### Potential mechanisms/explanations

Depression was associated with risk of MCVE among individuals regardless of binary SES category suggesting that mechanisms that link depression and CVD are present irrespective of socioeconomic backgrounds. Whilst individuals with depression and low SES were at particularly high risk of MCVE, the risk was additive for depression and each of low education and low income suggesting that there is little overlap in the underlying mechanisms of their association with CVD. In contrast, depression and area-based deprivation may share mechanisms that amplify each other in their effect on risk of CVD, perhaps more so among women than men. Individuals with severe depression may more often live in areas of high deprivation or the living environment itself influences the health of individuals, for example through social norms [36]. If one such social norm is not to seek help for depression, the association between depression and risk of CVD could be amplified by area-based deprivation.

### Implications of this study and future research

Our results suggest that individuals with depression and a lower socioeconomic background are at particularly high risk of MCVE. Monitoring of these individuals at high risk of adverse events needs to be encouraged, and timely diagnoses and effective treatments need to be offered to those affected by CVD. In the United Kingdom, the role of general practitioners is particularly important since they act as gate-keepers to health care services and are the first point of contact for individuals with depression and most other health problems. However, despite free access to most health care services for all residents in the United Kingdom, service provision and general practice funding is not aligned with clinical need, particularly in areas of high deprivation [37, 38]. Thus, it is important that the social gradient in risk of adverse events is not just highlighted in clinical guidelines but that resources and general practice funding is aligned with clinical need.

Since this is one of the first studies to report a potential synergistic effect of depression and area-based deprivation on risk of MCVE, a cautious interpretation of findings is warranted and it is important to establish whether the synergistic effect between depression and area-based deprivation is observed in other settings. Furthermore, this is the first study to comprehensively assess effect modification and interaction between depression and SES using different measures of SES. Future studies on the combined associations of depression and SES on risk of CVD should follow recommended reporting guidelines on effect modification and interaction [28] to facilitate comparison of results from different studies although international measures of SES are likely to differ.

## Conclusion

We found that depression was associated with increased risks of MCVE among those with low and high SES, irrespective of which measure of SES was used. Individuals with depression and a low SES were at particularly high risk of MCVE, particularly for the area-based deprivation measure. It is important that the social gradient in risk of adverse events is not just highlighted in clinical guidelines but that resources are allocated on the basis of clinical need to support implementation of guidelines.

## Data Availability

All bona fide researchers in academic, commercial, and charitable settings can apply to use the UK Biobank resource for health-related research in the public interest (www.ukbiobank.ac.uk/register-apply/).

## ACKNOWLEDGEMENTS

This research has been conducted using the UK Biobank Resource under application number 13797.

## Author contributions

RP, CAJ and SW conceived the study, RP performed the data analysis and wrote the draft manuscript, and CAJ and SW reviewed and edited the manuscript. RP is the guarantor of this work and takes responsibility for the contents of the article.

## Prior presentation

There has been no prior presentation of this work.

