## Supplementary material S1 for "The individual and combined associations of depression and socioeconomic status with risk of major cardiovascular events: a prospective cohort study"

**Supplementary Material S1: Ascertainment of covariates**

We ascertained covariates through self-report in the touchscreen questionnaire at baseline. We categorised ethnicity as white or other ethnicity. We used information smoking behaviour to identify participants as never, previous, or current smokers. We categorised alcohol intake as safe or risky drinking if men or women consumed ≤ 14 or > 14 units of alcohol per week, respectively (2). We assigned levels of physical activity as low, moderate or high, as recommended in published guidelines (3). We identified participants who consumed oily fish as at least once a week, less than once per week, and never. Sufficient daily fruit and vegetable intake was defined as at least five fruits or vegetables per day (4). We used information on self-reported illnesses of mother or father to determine a family history of stroke, heart disease, high blood pressure, and severe depression. We calculated body mass index ($kg/m^2$) based on measured values of height and weight at baseline, and categorised this as: underweight (<18.5 $kg/m^2$), normal weight (18.5 – 24.9 $kg/m^2$); overweight (25 – 29.9 $kg/m^2$); obese (30 – 34.9 $kg/m^2$); severely obese (35 – 39.9 $kg/m^2$); and morbidly obese (≥40 $kg/m^2$). We defined hypertension and high cholesterol levels as diagnosis and/or treatment, ascertained through self-report in the touchscreen questionnaire or nurse interview. Additionally, we identified participants with hypertension based on measured systolic and diastolic blood pressure at baseline (≥140/90 mmHg). We defined diabetes as one of self-reported diagnosis with or treatment for type 1 or 2 diabetes; or hospital record of type 1 or 2 diabetes at baseline (ICD 10: E10.X – E14.X). We identified participants with self-reported diabetes through information obtained in the touchscreen questionnaire and nurse interview. We defined treatment for diabetes as self-report of any medication listed in 'A10 Drugs used in diabetes' of the ATC/DDD Index 2018 [5].
