## Supplementary material S2 for "The individual and combined associations of depression and socioeconomic status with risk of major cardiovascular events: a prospective cohort study"

### Supplementary material S2: Baseline characteristics by missingness indicator

Table S1: Baseline characteristics for UK Biobank participants with and without complete information available

| | **Complete case** (n = 322,870) | **Missing** (n = 143,368) |
|---|---|---|
| **Men** | 155,450 (48.1) | 50,452 (35.2) |
| **Age** (median [IQR]) | 57.0 [49.0, 62.0] | 58.0 [50.0, 64.0] |
| **Ethnicity** | | |
| White | 308,540 (95.6) | 130,243 (90.8) |
| Other | 14,330 (4.4) | 10,596 (7.4) |
| **Highest educational attainment** | | |
| College or university degree | 122,446 (37.9) | 31,041 (21.7) |
| No college or university degree | 200,424 (62.1) | 103,181 (72.0) |
| **Area-based deprivation** | | |
| Less than UK avergage | 237,483 (73.6) | 97,142 (67.8) |
| More than UK average | 85,387 (26.4) | 45,650 (31.8) |
| **Income in £** | | |
| ≥ 31,000 | 181,783 (56.3) | 29,487 (20.6) |
| < 31,000 | 141,087 (43.7) | 43,699 (30.5) |
| **Body mass index** | | |
| Underweight (<18.5) | 1,635 (0.5) | 870 (0.6) |
| Normal weight (18.5 – 24.9) | 111,346 (34.5) | 44,215 (30.8) |
| Overweight (25 – 29.9) | 139,776 (43.3) | 57,003 (39.8) |
| Obese (30 – 34.9) | 52,046 (16.1) | 26,317 (18.4) |
| Severely obese (35 – 39.9) | 13,394 (4.1) | 8,541 (6.0) |
| Morbidly obese (≥40) | 4,673 (1.4) | 3,751 (2.6) |
| **Physical activity** | | |
| High | 122,345 (37.9) | 42,305 (29.5) |
| Moderate | 132,934 (41.2) | 48,842 (34.1) |
| Low | 67,591 (20.9) | 31,743 (22.1) |
| **Alcohol intake** | | |
| Safe drinking | 163,308 (50.6) | 42,634 (29.7) |
| Risky drinking | 159,562 (49.4) | 26,881 (18.7) |
| **Smoking status** | | |
| Never | 176,765 (54.7) | 82,463 (57.5) |
| Previous | 114,218 (35.4) | 42,294 (29.5) |
| Current | 31,887 (9.9) | 16,008 (11.2) |
| **Fruit and vegetable intake per day** | | |
| At least five a day | 95,672 (29.6) | 45,293 (31.6) |
| Less than five a day | 227,198 (70.4) | 96,538 (67.3) |
| **Oily fish intake** | | |
| At least once a week | 181,620 (56.3) | 75,406 (52.6) |
| Less than once a week | 108,987 (33.8) | 45,858 (32.0) |
| Never | 32,263 (10.0) | 18,456 (12.9) |
| **Depression** | 26,193 (8.1) | 14,456 (10.1) |
| **Hypertension** | 172,634 (53.5) | 80,555 (56.2) |
| **Diabetes** | 12,627 (3.9) | 7,952 (5.5) |
| **High cholesterol levels** | 45,985 (14.2) | 22,957 (16.0) |
| **Family history of CVD** | 223,813 (69.3) | 97,047 (67.7) |
| **Family history of depression** | 29,597 (9.2) | 11,705 (8.2) |

Data are number (%) unless otherwise indicated.
CVD: Cardiovascular disease; IQR: Interquartile range; UK: United Kingdom
