## Supplementary material S3 for "The individual and combined associations of depression and socioeconomic status with risk of major cardiovascular events: a prospective cohort study"

### Supplementary material S3: Complete case analysis

Table S2: Hazard ratios (95% CI) of the individuals effects of depression and socioeconomic status on risk of major cardiovascular events – complete cases

| | **Depression** (yes vs no) | **Educational attainment** (low vs high) | **Area-based deprivation** (high vs low) | **Income** (low vs high) |
|---|---|---|---|---|
| **Model 1** | 1.16 (1.05 - 1.28) | 1.51 (1.42 - 1.60) | 1.16 (1.09 - 1.24) | 1.81 (1.71 - 1.92) |
| **Model 2** | 1.41 (1.28 - 1.55) | 1.36 (1.28 - 1.45) | 1.27 (1.19 - 1.35) | 1.38 (1.30 - 1.46) |
| **Model 3** | 1.23 (1.11 - 1.35) | 1.19 (1.12 - 1.27) | 1.13 (1.06 - 1.20) | 1.21 (1.14 - 1.29) |
| **+ depression** | --- | 1.19 (1.12 - 1.27) | 1.12 (1.05 - 1.19) | 1.20 (1.13 - 1.28) |
| **+ education** | 1.22 (1.11 - 1.35) | --- | --- | --- |
| **+ deprivation** | 1.22 (1.11 - 1.35) | --- | --- | --- |
| **+ income** | 1.20 (1.09 - 1.32) | --- | --- | --- |

Data are hazard ratios (95% confidence intervals)
*Model 1: Depression, soecioeconomic factor, age, sex, ethnicity
†Model 2: Model 1 + body mass index, physical activity, alcohol intake, smoking, fruit and vegetable intake, oily fish intake, high cholesterol levels, hypertension, diabetes, family history of cardiovascular disease, and family history of depression

Table S3: Results of analyses on the combined association of depression and different measures of socioeconomic status on risk of major cardiovascular events – complete cases

| | | **Depression** | | | | HRs (95%CI) for depression by strata of SES measure |
|---|---|---|---|---|---|---|
| | | No | | Yes | | |
| | | n/N | HR (95%CI) | n/N | HR (95%CI) | |
| **Educational attainment** | High | 1,319 / 113,841 | ref. | 118 / 8,605 | 1.22 (1.01 – 1.48) | 1.22 (1.01 – 1.48) |
| | Low | 3,176 / 182,836 | 1.19 (1.11 – 1.27) | 340 / 17,588 | 1.46 (1.29 – 1.65) | 1.22 (1.09 – 1.37) |
| HRs (95%CI) for low education by depression status | | 1.19 (1.11 – 1.27) | | 1.19 (0.96 – 1.47) | | |
| **Area-based deprivation** | Low | 3,258 / 219,708 | ref. | 266 / 17,775 | 1.11 (0.98 – 1.26) | 1.11 (0.98 – 1.26) |
| | High | 1,237 / 76,969 | 1.09 (1.02 –1.17) | 192 / 8,418 | 1.56 (1.35 –1.81) | 1.43 (1.23 – 1.67) |
| HRs (95%CI) for high deprivation by depression status | | 1.09 (1.02 – 1.17) | | 1.41 (1.17 – 1.70) | | |
| **Income** | High | 1,950 / 170,746 | ref. | 131 / 11,037 | 1.16 (0.97 – 1.39) | 1.16 (0.97 – 1.39) |
| | Low | 2,545 / 125,931 | 1.20 (1.12 – 1.28) | 327 / 15,156 | 1.46 (1.29 – 1.64) | 1.22 (1.08 – 1.37) |
| HRs (95%CI) for low income by depression status | | 1.20 (1.12 – 1.28) | | 1.25 (1.02 – 1.54) | | |

HR are adjusted for age, sex, ethnicity, body mass index, physical activity, alcohol intake, smoking, fruit and vegetable intake, oily fish intake, high cholesterol levels, hypertension, diabetes, family history of cardiovascular disease, and family history of depression
CI = confidence interval; HR = hazard ratio; SES = socioeconomic status

Table S4: Measures of additive and multiplicative interaction between depression and different measures of socioeconomic status on risk of major cardiovascular events – complete cases

| | **Additive interaction** (RERI, 95% CI, p-value) | **Multiplicative interaction** (Ratio of HRs for SES measure within strata of depression, 95% C, p-value) |
|---|---|---|
| **Depression and education** | 0.04 (-0.24 – 0.32), p=0.384 | 1.00 (0.80 – 1.24), p=0.996 |
| **Depression and area-based deprivation** | 0.36 (0.10 – 0.63), p=0.004 | 1.29 (1.06 – 1.57), p=0.011 |
| **Depression and income** | 0.10 (-0.17 – 0.36), p=0.236 | 1.05 (0.85 – 1.29), p=0.677 |

Estimates (95% CI) are based on Cox proportional hazards models adjusted for age, sex, ethnicity, body mass index, physical activity, alcohol intake, smoking, fruit and vegetable intake, oily fish intake, high cholesterol levels, hypertension, diabetes, family history of cardiovascular disease, and family history of depression

CI = confidence interval; HR = hazard ratio; RERI = Relative Excess Risk due to Interaction; SES = socioeconomic status

Table S5: Hazard ratios (95% CI) of the individuals effects of depression and socioeconomic status on risk of major cardiovascular events – men – complete cases

| | **Depression** (yes vs no) | **Educational attainment** (low vs high) | **Area-based deprivation** (high vs low) | **Income** (low vs high) |
|---|---|---|---|---|
| **Model 1** | 1.43 (1.35 – 1.51) | 1.50 (1.40 – 1.61) | 1.12 (1.04 – 1.20) | 1.86 (1.74 – 1.99) |
| **Model 2** | 1.32 (1.16 – 1.50) | 1.36 (1.27 – 1.47) | 1.22 (1.13 – 1.31) | 1.36 (1.27 – 1.46) |
| **Model 3** | 1.17 (1.02 – 1.33) | 1.20 (1.12 – 1.30) | 1.10 (1.02 – 1.19) | 1.23 (1.14 – 1.32) |
| **+ depression** | —— | 1.20 (1.12 – 1.30) | 1.10 (1.02 – 1.18) | 1.22 (1.14 – 1.31) |
| **+ education** | 1.16 (1.02 – 1.33) | —— | —— | —— |
| **+ deprivation** | 1.16 (1.02 – 1.32) | —— | —— | —— |
| **+ income** | 1.13 (0.99 – 1.29) | —— | —— | —— |

Data are hazard ratios (95% confidence intervals)

*Model 1: Depression, soecioeconomic factor, age, sex, ethnicity

†Model 2: Model 1 + body mass index, physical activity, alcohol intake, smoking, fruit and vegetable intake, oily fish intake, high cholesterol levels, hypertension, diabetes, family history of cardiovascular disease, and family history of depression

Table S6: Results of analyses on the combined association of depression and different measures of socioeconomic status on risk of MCVE – men – complete cases

| | | **Depression** | | | | HRs (95%CI) for depression by strata of SES measure |
|---|---|---|---|---|---|---|
| | | No | | Yes | | |
| | | n/N | HR (95%CI) | n/N | HR (95%CI) | |
| **Educational attainment** | High | 797 / 57,090 | ref. | 69 / 3,133 | 1.16 (0.90 – 1.48) | 1.16 (0.90 – 1.48) |
| | Low | 2,284 / 89,589 | 1.20 (1.11 – 1.30) | 178 / 5,638 | 1.4 (1.19 – 1.65) | 1.17 (1.00 – 1.36) |
| HRs (95%CI) for low education by depression status | | 1.20 (1.11 – 1.30) | | 1.21 (0.92 – 1.60) | | |

| | | | | | | |
|---|---|---|---|---|---|---|
| **Area-based deprivation** | Low | 2,382 / 108,749 | ref. | 146 / 5,728 | 1.07 (0.91 – 1.27) | 1.07 (0.91 – 1.27) |
| | High | 881 / 37,930 | 1.08 (1.00 – 1.17) | 101 / 3,043 | 1.41 (1.16 – 1.73) | 1.31 (1.07 – 1.61) |
| HRs (95%CI) for high deprivation by depression status | | 1.08 (1.00 – 1.17) | | 1.32 (1.02 – 1.70) | | |
| **Income** | High | 1,506 / 89,007 | ref. | 71 / 3,825 | 1.02 (0.81 – 1.30) | 1.02 (0.81 – 1.30) |
| | Low | 1,757 / 57,672 | 1.21 (1.12 – 1.30) | 176 / 4,946 | 1.43 (1.22 – 1.68) | 1.19 (1.01 – 1.39) |
| HRs (95%CI) for low income by depression status | | 1.21 (1.12 – 1.30) | | 1.40 (1.06 – 1.85) | | |

HR are adjusted for age, sex, ethnicity, body mass index, physical activity, alcohol intake, smoking, fruit and vegetable intake, oily fish intake, high cholesterol levels, hypertension, diabetes, family history of cardiovascular disease, and family history of depression
CI = confidence interval; HR = hazard ratio; SES = socioeconomic status

Table S7: Measures of additive and multiplicative interaction between depression and different measures of socioeconomic status on risk of MCVE - men – complete cases

| | **Additive interaction** (RERI, 95% CI, p-value) | **Multiplicative interaction** (Ratio of HRs for SES measure within strata of depression, 95% C, p-value) |
|---|---|---|
| **Depression and education** | 0.05 (-0.31 – 0.40), p=0.401 | 1.01 (0.76 – 1.35), p=0.947 |
| **Depression and area-based deprivation** | 0.26 (-0.07 – 0.60), p=0.063 | 1.22 (0.94 – 1.59), p=0.139 |
| **Depression and income** | 0.20 (-0.12 – 0.53), p=0.111 | 1.16 (0.87 – 1.54), p=0.304 |

Estimates (95% CI) are based on Cox proportional hazards models adjusted for age, sex, ethnicity, body mass index, physical activity, alcohol intake, smoking, fruit and vegetable intake, oily fish intake, high cholesterol levels, hypertension, diabetes, family history of cardiovascular disease, and family history of depression
CI = confidence interval; HR = hazard ratio; RERI = Relative Excess Risk due to Interaction; SES = socioeconomic status

Table S8: Hazard ratios (95% CI) of the individuals effects of depression and socioeconomic status on risk of major cardiovascular events – women – complete cases

| | **Depression** (yes vs no) | **Educational attainment** (low vs high) | **Area-based deprivation** (high vs low) | **Income** (low vs high) |
|---|---|---|---|---|
| **Model 1** | 1.48 (1.28 – 1.71) | 1.62 (1.44 – 1.82) | 1.28 (1.15 – 1.43) | 2.14 (1.92 – 2.39) |
| **Model 2** | 1.53 (1.32 – 1.77) | 1.34 (1.20 – 1.51) | 1.40 (1.25 – 1.56) | 1.40 (1.25 – 1.57) |
| **Model 3** | 1.30 (1.12 – 1.51) | 1.15 (1.02 – 1.30) | 1.19 (1.06 – 1.33) | 1.18 (1.05 – 1.33) |
| **+ depression** | — | 1.15 (1.02 – 1.29) | 1.18 (1.05 – 1.33) | 1.16 (1.03 – 1.31) |
| **+ education** | 1.30 (1.12 – 1.51) | — | — | — |
| **+ deprivation** | 1.29 (1.12 – 1.50) | — | — | — |
| **+ income** | 1.28 (1.11 – 1.49) | — | — | — |

Data are hazard ratios (95% confidence intervals)
*Model 1: Depression, soecioeconomic factor, age, sex, ethnicity

†Model 2: Model 1 + body mass index, physical activity, alcohol intake, smoking, fruit and vegetable intake, oily fish intake, high cholesterol levels, hypertension, diabetes, family history of cardiovascular disease, and family history of depression

Table S9: Results of analyses on the combined association of depression and different measures of socioeconomic status on risk of MCVE – women – complete cases

| | | Depression | | | | HRs (95%CI) for depression by strata of SES measure |
|---|---|---|---|---|---|---|
| | | No | | Yes | | |
| | | n/N | HR (95%CI) | n/N | HR (95%CI) | |
| **Educational attainment** | High | 340 / 56,751 | ref. | 49 / 5,472 | 1.34 (0.99 – 1.81) | 1.34 (0.99 – 1.81) |
| | Low | 892 / 93,247 | 1.15 (1.02 – 1.31) | 162 / 11,950 | 1.48 (1.22 – 1.80) | 1.28 (1.08 – 1.52) |
| HRs (95%CI) for low education by depression status | | 1.15 (1.02 – 1.31) | | 1.10 (0.80 – 1.52) | | |
| **Area-based deprivation** | Low | 876 / 110,959 | ref. | 120 / 12,047 | 1.16 (0.96 – 1.41) | 1.16 (0.96 – 1.41) |
| | High | 356 / 39,039 | 1.13 (0.99 – 1.28) | 91 / 5,375 | 1.75 (1.40 – 2.18) | 1.55 (1.23 – 1.96) |
| HRs (95%CI) for high deprivation by depression status | | 1.13 (0.99 – 1.28) | | 1.51 (1.14 – 1.98) | | |
| **Income** | High | 444 / 81,739 | ref. | 60 / 7,212 | 1.43 (1.09 – 1.88) | 1.43 (1.09 – 1.88) |
| | Low | 788 / 68,259 | 1.19 (1.05 – 1.34) | 151 / 10,210 | 1.46 (1.20 – 1.77) | 1.23 (1.03 – 1.47) |
| HRs (95%CI) for low income by depression status | | 1.19 (1.05 – 1.34) | | 1.02 (0.75 – 1.38) | | |

HR are adjusted for age, sex, ethnicity, body mass index, physical activity, alcohol intake, smoking, fruit and vegetable intake, oily fish intake, high cholesterol levels, hypertension, diabetes, family history of cardiovascular disease, and family history of depression

CI = confidence interval; HR = hazard ratio; SES = socioeconomic status

Table S10: Measures of additive and multiplicative interaction between depression and different measures of socioeconomic status on risk of MCVE - women – complete cases

| | **Additive interaction** (RERI, 95% CI, p-value) | **Multiplicative interaction** (Ratio of HRs for SES measure within strata of depression, 95% C, p-value) |
|---|---|---|
| **Depression and education** | -0.01 (-0.47 – 0.45), p=0.524 | 0.96 (0.68 – 1.35), p=0.802 |
| **Depression and area-based deprivation** | 0.46 (0.03 – 0.89), p=0.019 | 1.34 (0.99 – 1.80), p=0.058 |
| **Depression and income** | -0.16 (-0.61 – 0.29), p=0.754 | 0.86 (0.62 – 1.18), p=0.355 |

Estimates (95% CI) are based on Cox proportional hazards models adjusted for age, sex, ethnicity, body mass index, physical activity, alcohol intake, smoking, fruit and vegetable intake, oily fish intake, high cholesterol levels, hypertension, diabetes, family history of cardiovascular disease, and family history of depression

CI = confidence interval; HR = hazard ratio; RERI = Relative Excess Risk due to Interaction; SES = socioeconomic status

Table S11: Hazard ratios (95% CI) of the individuals effects of depression and socioeconomic status on risk of stroke – complete cases

| | **Depression** (yes vs no) | **Educational attainment** (low vs high) | **Area–based deprivation** (high vs low) | **Income** (low vs high) |
|---|---|---|---|---|
| **Model 1** | 1.21 (1.04 – 1.40) | 1.45 (1.32 – 1.60) | 1.27 (1.16 – 1.40) | 2.00 (1.83 – 2.19) |
| **Model 2** | 1.37 (1.18 – 1.59) | 1.27 (1.15 – 1.39) | 1.42 (1.29 – 1.56) | 1.40 (1.27 – 1.54) |
| **Model 3** | 1.24 (1.06 – 1.44) | 1.14 (1.03 – 1.26) | 1.28 (1.16 – 1.41) | 1.27 (1.15 – 1.40) |
| **+ depression** | — | 1.14 (1.03 – 1.25) | 1.28 (1.16 – 1.41) | 1.26 (1.14 – 1.39) |
| **+ education** | 1.24 (1.06 – 1.44) | — | — | — |
| **+ deprivation** | 1.22 (1.05 – 1.43) | — | — | — |
| **+ income** | 1.21 (1.04 – 1.41) | — | — | — |

Data are hazard ratios (95% confidence intervals)
*Model 1: Depression, soecioeconomic factor, age, sex, ethnicity
†Model 2: Model 1 + body mass index, physical activity, alcohol intake, smoking, fruit and vegetable intake, oily fish intake, high cholesterol levels, hypertension, diabetes, family history of cardiovascular disease, and family history of depression

Table S12: Results of analyses on the combined association of depression and different measures of socioeconomic status on risk of stroke – complete cases

| | | **Depression** | | | | HRs (95%CI) for depression by strata of SES measure |
|---|---|---|---|---|---|---|
| | | No | | Yes | | |
| | | n/N | HR (95%CI) | n/N | HR (95%CI) | |
| **Educational attainment** | High | 535 / 113,841 | ref. | 49 / 8,605 | 1.22 (0.91 – 1.64) | 1.22 (0.91 –1.64) |
| | Low | 1,237 / 182,836 | 1.13 (1.02 – 1.26) | 139 / 17,588 | 1.41 (1.16 – 1.70) | 1.24 (1.04 – 1.48) |
| HRs (95%CI) for low education by depression status | | | 1.13 (1.02, 1.26) | | 1.15 (0.83 – 1.59) | |
| **Area-based deprivation** | Low | 1,253 / 219,708 | ref. | 104 / 17,775 | 1.08 (0.89 – 1.33) | 1.08 (0.89 – 1.33) |
| | High | 519 / 76,969 | 1.23 (1.11 – 1.37) | 84 / 8,418 | 1.82 (1.45 – 2.28) | 1.47 (1.17 –1.86) |
| HRs (95%CI) for high deprivation by depression status | | | 1.23 (1.11 – 1.37) | | 1.68 (1.26 – 2.24) | |
| **Income** | High | 718 / 170,746 | ref. | 58 / 11,037 | 1.35 (1.03 – 1.76) | 1.35 (1.03 – 1.76) |
| | Low | 1,054 / 125,931 | 1.27 (1.15 – 1.41) | 130 / 15,156 | 1.47 (1.21 – 1.78) | 1.15 (0.96 – 1.38) |
| HRs (95%CI) for low income by depression status | | | 1.27 (1.15 – 1.41) | | 1.09 (0.80 – 1.48) | |

HR are adjusted for age, sex, ethnicity, body mass index, physical activity, alcohol intake, smoking, fruit and vegetable intake, oily fish intake, high cholesterol levels, hypertension, diabetes, family history of cardiovascular disease, and family history of depression
CI = confidence interval; HR = hazard ratio; SES = socioeconomic status

Table S13: Measures of additive and multiplicative interaction between depression and different measures of socioeconomic status on risk of stroke – complete cases

| | **Additive interaction** (RERI, 95% CI, p-value) | **Multiplicative interaction** (Ratio of HRs for SES measure within strata of depression, 95% C, p-value) |
|---|---|---|
| **Depression and education** | 0.05 (-0.38 – 0.48), p=0.411 | 1.01 (0.72 – 1.43), p=0.937 |
| **Depression and area-based deprivation** | 0.50 (0.04 – 0.96), p=0.016 | 1.36 (1.00 – 1.84), p=0.049 |
| **Depression and income** | -0.16 (-0.60 – 0.29), p=0.754 | 0.85 (0.62 – 1.18), p=0.340 |

Estimates (95% CI) are based on Cox proportional hazards models adjusted for age, sex, ethnicity, body mass index, physical activity, alcohol intake, smoking, fruit and vegetable intake, oily fish intake, high cholesterol levels, hypertension, diabetes, family history of cardiovascular disease, and family history of depression
CI = confidence interval; HR = hazard ratio; RERI = Relative Excess Risk due to Interaction; SES = socioeconomic status

Table S14: Hazard ratios (95% CI) of the individuals effects of depression and socioeconomic status on risk of myocardial infarction – complete cases

| | **Depression** (yes vs no) | **Educational attainment** (low vs high) | **Area-based deprivation** (high vs low) | **Income** (low vs high) |
|---|---|---|---|---|
| **Model 1** | 1.14 (1.01 – 1.29) | 1.56 (1.44 – 1.68) | 1.11 (1.02 – 1.20) | 1.72 (1.61 – 1.85) |
| **Model 2** | 1.44 (1.27 – 1.63) | 1.43 (1.32 – 1.55) | 1.20 (1.11 – 1.30) | 1.37 (1.27 – 1.48) |
| **Model 3** | 1.23 (1.08 – 1.39) | 1.23 (1.13 – 1.33) | 1.05 (0.97 – 1.13) | 1.19 (1.10 – 1.28) |
| **+ depression** | —— | 1.23 (1.13 – 1.33) | 1.04 (0.96 – 1.13) | 1.18 (1.09 – 1.27) |
| **+ education** | 1.22 (1.08 – 1.38) | —— | —— | —— |
| **+ deprivation** | 1.22 (1.08 – 1.39) | —— | —— | —— |
| **+ income** | 1.20 (1.06 – 1.36) | —— | —— | —— |

Data are hazard ratios (95% confidence intervals)
*Model 1: Depression, soecioeconomic factor, age, sex, ethnicity
†Model 2: Model 1 + body mass index, physical activity, alcohol intake, smoking, fruit and vegetable intake, oily fish intake, high cholesterol levels, hypertension, diabetes, family history of cardiovascular disease, and family history of depression

Table S15: Results of analyses on the combined association of depression and different measures of socioeconomic status on risk of myocardial infarction – complete cases

| | | **Depression** | | | | HRs (95%CI) for depression by strata of SES measure |
|---|---|---|---|---|---|---|
| | | No | | Yes | | |
| | | n/N | HR (95%CI) | n/N | HR (95%CI) | |
| **Educational attainment** | High | 804 / 113,841 | ref. | 74 / 8,605 | 1.28 (1.01 – 1.62) | 1.28 (1.01 – 1.62) |
| | Low | 2,010 / 182,836 | 1.23 (1.14 – 1.34) | 207 / 17,588 | 1.49 (1.27 – 1.74) | 1.20 (1.04 – 1.39) |
| HRs (95%CI) for low education by depression status | | | 1.23 (1.14 – 1.34) | | 1.16 (0.89 – 1.52) | |
| **Area-based deprivation** | Low | 2,066 / 219,708 | ref. | 168 / 17,775 | 1.13 (0.96 – 1.32) | 1.13 (0.96 – 1.32) |

| | | | | | | |
|---|---|---|---|---|---|---|
| | High | 748 / 76,969 | 1.02 (0.93 – 1.11) | 113 / 8,418 | 1.43 (1.18 – 1.73) | 1.40 (1.15 – 1.71) |
| HRs (95%CI) for high deprivation by depression status | | | 1.02 (0.93 – 1.11) | | 1.26 (0.99 – 1.60) | |
| **Income** | High | 1,262 / 170,746 | ref. | 75 / 11,037 | 1.05 (0.83 – 1.33) | 1.05 (0.83 – 1.33) |
| | Low | 1,552 / 125,931 | 1.16 (1.07 – 1.26) | 206 / 15,156 | 1.47 (1.27 – 1.71) | 1.27 (1.09 – 1.47) |
| HRs (95%CI) for low income by depression status | | | 1.16 (1.07 – 1.26) | | 1.40 (1.07 – 1.82) | |

HR are adjusted for age, sex, ethnicity, body mass index, physical activity, alcohol intake, smoking, fruit and vegetable intake, oily fish intake, high cholesterol levels, hypertension, diabetes, family history of cardiovascular disease, and family history of depression
CI = confidence interval; HR = hazard ratio; SES = socioeconomic status

Table S16: Measures of additive and multiplicative interaction between depression and different measures of socioeconomic status on risk of myocardial infarction – complete cases

| | **Additive interaction** (RERI, 95% CI, p-value) | **Multiplicative interaction** (Ratio of HRs for SES measure within strata of depression, 95% C, p-value) |
|---|---|---|
| **Depression and education** | -0.03 (-0.39 – 0.34), p=0.555 | 0.94 (0.71 – 1.24), p=0.675 |
| **Depression and area-based deprivation** | 0.28 (-0.05 – 0.6), p=0.047 | 1.24 (0.96 – 1.60), p=0.097 |
| **Depression and income** | 0.26 (-0.06 – 0.58), p=0.057 | 1.20 (0.92 – 1.59), p=0.185 |

Estimates (95% CI) are based on Cox proportional hazards models adjusted for age, sex, ethnicity, body mass index, physical activity, alcohol intake, smoking, fruit and vegetable intake, oily fish intake, high cholesterol levels, hypertension, diabetes, family history of cardiovascular disease, and family history of depression
CI = confidence interval; HR = hazard ratio; RERI = Relative Excess Risk due to Interaction; SES = socioeconomic status

Table S17: Hazard ratios (95% CI) of the individuals effects of depression and socioeconomic status on risk of deaths from causes other than stroke or myocardial infarction – complete cases

| | **Depression** (yes vs no) | **Educational attainment** (low vs high) | **Area-based deprivation** (high vs low) | **Income** (low vs high) |
|---|---|---|---|---|
| **Model 1** | 1.34 (1.23 – 1.46) | 1.44 (1.36 – 1.52) | 1.30 (1.23 – 1.38) | 2.19 (2.07 – 2.31) |
| **Model 2** | 1.51 (1.39 – 1.65) | 1.26 (1.19 – 1.33) | 1.46 (1.38 – 1.55) | 1.58 (1.49 – 1.67) |
| **Model 3** | 1.34 (1.23 – 1.47) | 1.13 (1.07 – 1.20) | 1.28 (1.21 – 1.35) | 1.43 (1.35 – 1.51) |
| **+ depression** | — | 1.13 (1.06 – 1.20) | 1.27 (1.20 – 1.35) | 1.41 (1.33 – 1.49) |
| **+ education** | 1.34 (1.23 – 1.46) | — | — | — |
| **+ deprivation** | 1.33 (1.22 – 1.45) | — | — | — |
| **+ income** | 1.29 (1.18 – 1.41) | — | — | — |

Data are hazard ratios (95% confidence intervals)
*Model 1: Depression, soecioeconomic factor, age, sex, ethnicity

†Model 2: Model 1 + body mass index, physical activity, alcohol intake, smoking, fruit and vegetable intake, oily fish intake, high cholesterol levels, hypertension, diabetes, family history of cardiovascular disease, and family history of depression

Table S18: Results of analyses on the combined association of depression and different measures of socioeconomic status on risk of deaths from causes other than stroke or myocardial infarction – complete cases

| | | Depression | | | | HRs (95%CI) for depression by strata of SES measure |
|---|---|---|---|---|---|---|
| | | No | | Yes | | |
| | | n/N | HR (95%CI) | n/N | HR (95%CI) | |
| **Educational attainment** | High | 1,528 / 113,841 | ref. | 170 / 8,605 | 1.47 (1.26 – 1.73) | 1.47 (1.26 – 1.73) |
| | Low | 3,533 / 182,836 | 1.14 (1.08 – 1.22) | 424 / 17,588 | 1.48 (1.33 – 1.65) | 1.29 (1.17 – 1.43) |
| HRs (95%CI) for low education by depression status | | 1.14 (1.08 – 1.22) | | 1.00 (0.84 – 1.20) | | |
| **Area-based deprivation** | Low | 3,528 / 219,708 | ref. | 363 / 17,775 | 1.33 (1.19 – 1.48) | 1.33 (1.19 – 1.48) |
| | High | 1,533 / 76,969 | 1.27 (1.19 – 1.35) | 231 / 8,418 | 1.69 (1.47 – 1.93) | 1.33 (1.16 – 1.53) |
| HRs (95%CI) for high deprivation by depression status | | 1.27 (1.19 – 1.35) | | 1.27 (1.08 – 1.50) | | |
| **Income** | High | 1,963 / 170,746 | ref. | 159 / 11,037 | 1.33 (1.13 – 1.56) | 1.33 (1.13 – 1.56) |
| | Low | 3,098 / 125,931 | 1.41 (1.33 – 1.50) | 435 / 15,156 | 1.81 (1.62 – 2.01) | 1.28 (1.15 – 1.42) |
| HRs (95%CI) for low income by depression status | | 1.41 (1.33 – 1.50) | | 1.36 (1.13 – 1.63) | | |

HR are adjusted for age, sex, ethnicity, body mass index, physical activity, alcohol intake, smoking, fruit and vegetable intake, oily fish intake, high cholesterol levels, hypertension, diabetes, family history of cardiovascular disease, and family history of depression
CI = confidence interval; HR = hazard ratio; SES = socioeconomic status

Table S19: Measures of additive and multiplicative interaction between depression and different measures of socioeconomic status on risk of deaths from causes other than stroke or myocardial infarction – complete cases

| | **Additive interaction** (RERI, 95% CI, p-value) | **Multiplicative interaction** (Ratio of HRs for SES measure within strata of depression, 95% C, p-value) |
|---|---|---|
| **Depression and education** | -0.14 (-0.41 – 0.13), p=0.840 | 0.88 (0.73 – 1.06), p=0.175 |
| **Depression and area-based deprivation** | 0.09 (-0.17 – 0.36), p=0.250 | 1.00 (0.84 – 1.19), p=0.980 |
| **Depression and income** | 0.06 (-0.21 – 0.34), p=0.326 | 0.96 (0.79 – 1.16), p=0.683 |

Estimates (95% CI) are based on Cox proportional hazards models adjusted for age, sex, ethnicity, body mass index, physical activity, alcohol intake, smoking, fruit and vegetable intake, oily fish intake, high cholesterol levels, hypertension, diabetes, family history of cardiovascular disease, and family history of depression
CI = confidence interval; HR = hazard ratio; RERI = Relative Excess Risk due to Interaction; SES = socioeconomic status
