## Supplementary material S6 for "The individual and combined associations of depression and socioeconomic status with risk of major cardiovascular events: a prospective cohort study"

### Supplementary material S6: Sensitivity analyses – competing risk analysis

Table S32: Hazard ratios (95% CI) of the individuals effects of depression and socioeconomic status on risk of deaths from causes other than stroke or myocardial infarction

| | **Depression** (yes vs no) | **Educational attainment** (low vs high) | **Area-based deprivation** (high vs low) | **Income** (low vs high) |
|---|---|---|---|---|
| **Model 1** | 1.39 (1.30 – 1.48) | 1.46 (1.39 – 1.53) | 1.32 (1.26 – 1.38) | 2.02 (1.93 – 2.12) |
| **Model 2** | 1.57 (1.47 – 1.67) | 1.27 (1.21 – 1.34) | 1.49 (1.43 – 1.56) | 1.49 (1.42 – 1.56) |
| **Model 3** | 1.39 (1.30 – 1.48) | 1.13 (1.08 – 1.19) | 1.28 (1.23 – 1.34) | 1.32 (1.25 – 1.39) |
| **+ depression** | — | 1.13 (1.08 – 1.19) | 1.27 (1.22 – 1.33) | 1.30 (1.24 – 1.37) |
| **+ education** | 1.38 (1.30 – 1.48) | — | — | — |
| **+ deprivation** | 1.37 (1.28 – 1.46) | — | — | — |
| **+ income** | 1.35 (1.26 – 1.44) | — | — | — |

Data are hazard ratios (95% confidence intervals)
*Model 1: Depression, soecioeconomic factor, age, sex, ethnicity
†Model 2: Model 1 + body mass index, physical activity, alcohol intake, smoking, fruit and vegetable intake, oily fish intake, high cholesterol levels, hypertension, diabetes, family history of cardiovascular disease, and family history of depression

Table S33: Results of analyses on the combined association of depression and different measures of socioeconomic status on risk of deaths from causes other than stroke or myocardial infarction

| | | **Depression** | | | | HRs (95%CI) for depression by strata of SES measure |
|---|---|---|---|---|---|---|
| | | No | | Yes | | |
| | | n/N | HR (95%CI) | n/N | HR (95%CI) | |
| **Educational attainment** | High | 2,024 / 142,113 | ref. | 232 / 11,374 | 1.44 (1.26 – 1.65) | 1.44 (1.26 – 1.65) |
| | Low | 5,714 / 275,058 | 1.14 (1.08 – 1.20) | 1,054 / 37,693 | 1.45 (1.34 – 1.57) | 1.38 (1.28 – 1.49) |
| HRs (95%CI) for low education by depression status | | | 1.14 (1.08 – 1.20) | | 1.11 (0.96 - 1.30) | |
| **Area-based deprivation** | Low | 5,389 / 308,148 | ref. | 603 / 26,477 | 1.34 (1.23 – 1.46) | 1.34 (1.23 – 1.46) |
| | High | 2,576 / 116,937 | 1.27 (1.21 – 1.33) | 456 / 14,676 | 1.75 (1.58 – 1.93) | 1.43 (1.29 – 1.59) |
| HRs (95%CI) for high deprivation by depression status | | | 1.27 (1.21 – 1.33) | | 1.32 (1.16 - 1.50) | |
| **Income** | High | 2,313 / 197,908 | ref. | 201 / 13,362 | 1.36 (1.18 – 1.57) | 1.36 (1.18 – 1.57) |
| | Low | 4,138 / 163,848 | 1.42 (1.35 – 1.50) | 2,372 / 91,120 | 1.56 (1.46 – 1.65) | 1.34 (1.24 – 1.45) |
| HRs (95%CI) for low income by depression status | | | 1.42 (1.35 – 1.50) | | 1.32 (1.13 - 1.56) | |

HR are adjusted for age, sex, ethnicity, body mass index, physical activity, alcohol intake, smoking, fruit and vegetable intake, oily fish intake, high cholesterol levels, hypertension, diabetes, family history of cardiovascular disease, and family history of depression
CI = confidence interval; HR = hazard ratio; SES = socioeconomic status

Table S34: Measures of additive and multiplicative interaction between depression and different measures of socioeconomic status on risk of deaths from causes other than stroke or myocardial infarction

| | **Additive interaction** (RERI, 95% CI, p-value) | **Multiplicative interaction** (Ratio of HRs for SES measure within strata of depression, 95% C, p-value) |
|---|---|---|
| **Depression and education** | -0.13 (-0.34 – 0.08), p = 0.884 | 0.95 (0.81 – 1.10), p = 0.478 |
| **Depression and area-based deprivation** | 0.14 (-0.06 – 0.35), p = 0.080 | 1.05 (0.92 – 1.20), p = 0.487 |
| **Depression and income** | -0.23 (-0.44 – -0.02), p = 0.983 | 0.98 (0.83 – 1.15), p = 0.776 |

Estimates (95% CI) are based on Cox proportional hazards models adjusted for age, sex, ethnicity, body mass index, physical activity, alcohol intake, smoking, fruit and vegetable intake, oily fish intake, high cholesterol levels, hypertension, diabetes, family history of cardiovascular disease, and family history of depression
CI = confidence interval; HR = hazard ratio; RERI = Relative Excess Risk due to Interaction; SES = socioeconomic status
